# Urinary extracellular vesicles in healthy individuals: positive correlation between podocyte and tubular vesicles independent of kidney function

**DOI:** 10.1101/2024.08.04.24311469

**Authors:** Liang Wu, Carla C. Baan, Derek Reijerkerk, Dennis A. Hesselink, Karin Boer

## Abstract

Urinary extracellular vesicles (uEVs) are promising non-invasive biomarkers for assessing renal physiology and disease. Focusing specifically on kidney-derived uEVs (kd-uEVs) rather than the overall uEV population may offer a more precise insight into kidney health. However, research distinguishing single kd-uEVs from various nephron segments and their relationship with kidney biology remains limited. Imaging flow cytometry (IFCM) can identify single kd-uEVs by labeling them with CD63 (a uEV marker) in combination with PODXL (a podocyte marker) or AQP2 (a tubular marker). This study investigated the correlations between CD63+ AQP2+ or PODXL+ kd-uEVs and kidney function. CD63+ AQP2+ and CD63+ PODXL+ uEVs were detected in urine compared to negative controls, including urine with detergent treatment or isotype staining and reagent-containing PBS. While no significant association was found between CD63+ AQP2+ or PODXL+ uEV concentration and kidney function, a significant correlation was observed between AQP2+ and PODXL+ uEV concentrations (Rho = 0.789, p < 0.001). This correlation could be explained by the colocalization of AQP2 and PODXL on CD63+ uEVs. In conclusion, our study is the first to demonstrate the colocalization of podocyte and tubular proteins on uEVs.

## Introduction

Urinary extracellular vesicles (uEVs) are potential non-invasive biomarkers of renal physiology and disease [1, 2]. While the kidney is the primary source, uEVs also derive from the lower urinary tract and the bloodstream [1]. Rather than measuring the overall uEV population, focusing on kidney-derived uEVs (kd-uEVs) may provide a more accurate picture of kidney health. Proteomic analysis of the entire uEVs demonstrated that podocalyxin (PODXL; podocyte marker) and aquaporin 2 (AQP2; tubule/collecting duct marker) are the most abundant kidney-specific proteins [3]. However, research on the discrimination of single kd-uEVs from different nephron segments and their correlation with kidney biology remains limited. Imaging flow cytometry (IFCM) can identify single kd-uEVs by labeling them with CD63 (uEV marker), combined with PODXL or AQP2 [4, 5]. This study aimed to explore the correlations between CD63+ AQP2+ or PODXL+ kd-uEVs and sex, age, and kidney function.

## Methods and Materials

### Patients and urine collection

Spot urine samples were collected from 30 healthy kidney transplant donors at the Erasmus MC, Rotterdam, The Netherlands, between August 2018 and August 2021 (clinical parameters are summarized in **Supplementary Table S1**). Urine processing protocols, including centrifugation and addition of a proteinase inhibitor to the supernatant, were described previously [5].

### Isolation-free labeling of uEVs for IFCM

For the labeling of kidney-derived uEVs, 112 μL of urine sample underwent staining with 4 μL of a 30×-diluted anti-CD63 monoclonal antibody conjugated with allophycocyanin (CD63-APC, 200 μg/mL, clone H5C6, Biolegend, San Diego, CA, USA), alongside 4 μL of a kidney-specific antibody, in a 96-well plate placed in darkness overnight at room temperature. The kidney-specific antibody, either a 15×-diluted anti-AQP2 conjugated with Alexa Fluor 488 (AQP2-Alexa488, 200 μg/mL, clone E-2) or a 30×-diluted PODXL-Alexa488 (200 μg/mL, clone 3D3), was obtained from Santa Cruz Animal Health (Dallas, TX, USA).

To explore the colocalization of AQP2 and PODXL on CD63+ uEVs, a triple-staining protocol was executed for IFCM. Herein, 108 μL of urine sample was stained with 4 μL of a 15×-diluted anti-CD63-eFluor450 (200 μg/mL, clone H5C6, ThermoFisher Scientific, Waltham, MA, USA), 4 μL of a 15×-diluted AQP2-Alexa488, and 4 μL of a 20×-diluted PODXL-Alexa647 in darkness overnight at room temperature.

Before staining, all antibodies were centrifuged at 16,000 g for 10 min. Each stained urine sample was triplicated within the plate, with subsequent analysis employing the median value of the detected uEV concentration. Non-biological background signals, such as antibody aggregates, were identified based on detergent treatment, which lyses phospholipid and lipid particles. Following each IFCM measurement, all labeled samples were incubated with 5 μL of TritonX-100 (final concentration of 0.05%) at room temperature for 30 min before re-measurement. Staining specificity was verified using isotypes conjugating with the same fluorophores and at the same concentrations (**Supplementary Table S2**).

### Data acquisition and gating strategy for IFCM

The acquisition of Imaging Flow Cytometry (IFCM) data was conducted using an ImageStreamX Mark II instrument (ISx; Cytek Biosciences, Fremont, CA, USA) installed with INSPIRE® software (version 200.1.0.765; Cytek Biosciences), following established protocols for analyzing unprocessed urine and plasma [5, 6]. The INSPIRE® settings were configured as follows: a flow speed velocity of 40 mm/s, a flow core diameter of 6 μm, 60× magnification, and laser wavelengths of 405 nm (channel 01), 488 nm (channel 02), 642 nm (channel 05), and 785 nm for side scatter (SSC) excitation (channel 06). Additionally, bright-field imaging was conducted using channel 04. Fluorescence signals from eFluor450, Alexa488, and APC (or Alexa647) were detected in channels 01, 02, and 05, respectively.

Quantification of samples was performed using Amnis IDEAS software (version 6.2; Cytek Biosciences) [5, 6]. To ensure the quantification of individual urinary extracellular vesicles, a specific gating strategy was employed as previously described [6]. In brief: 1) particles exhibiting SSC intensities ≤ 5279 a.u., corresponding to EVs ≤ 1200 nm based on calibration, were selected; 2) only singlets were included, while objects with multiple fluorescent spots were excluded; 3) due to the presence of auto-fluorescence (A-F) particles in unprocessed urine, which exhibit positivity in all channels even without antibody staining and cannot be removed by detergent [6], A-F particles showing positivity in an empty channel 03 (no antibody) were excluded from the analysis; 4) gates were set up to identify single, double, or triple-positive uEVs based on unstained, isotype-stained, and single-stained control samples.

### Immunohistochemistry

The tissue specificity of anti-AQP2 (clone E2, Santa Cruz) and anti-PODXL (clone 3D3, Santa Cruz) antibodies was assessed via immunohistochemistry experiments conducted on formalin-fixed paraffin-embedded (FFPE) sections obtained from residual peritumoral tissue from nephrectomies performed for oncological purposes (**Supplementary Fig. S1**). Immunohistochemistry Chromogenic multiplex (cmIHC) analysis was conducted using an automated cmIHC system, specifically the Ventana Benchmark DiscoverAy ULTRA (Ventana Medical Systems Inc., Oro Valley, AZ, USA).

In summary, 4 μm thick tissue sections were stained following deparaffinization and heat-induced antigen retrieval with CC1 (#950-224, Ventana) for 32 minutes. A 4000× dilution of anti-podocalyxin ligand 1 was incubated at 37°C for 32 minutes, followed by treatment with omnimap anti-mouse HRP (#760-4310, Ventana) and detection with purple (#760-299, Ventana) for 32 minutes. An antibody denaturation step was then performed with CC2 (#950-123, Ventana) at 100°C for 20 minutes. Subsequently, incubation with a 100× dilution of anti-AQP2 was conducted at 37°C for 32 minutes, followed by mouse-NP and anti-NP-AP (#760-4816 and #760-4827, Ventana) and detection with the Yellow AP kit (#760-239, Ventana). Counterstaining was performed with hematoxylin II for 4 minutes, followed by a blue coloring reagent for an additional 4 minutes, as per the manufacturer’s instructions (Ventana). Finally, slides were scanned using the Hamamatsu Nanozoomer (Hamamatsu Photonics, Hamamatsu, Japan).

Reagent-containing phosphate-buffered saline (PBS), detergent-treated urine, and isotype-stained urine were used to validate the uEV measurement [1]. As recommended by the International Society for Extracellular Vesicles [1], urine dilution was normalized by calculating the concentration ratio of uEVs to urinary creatinine (UCr) measured with CRE2U, ACN 8152 (Roche Diagnostics Nederland BV).

### Statistical analysis

Data were presented as median [quartile 1 – quartile 3; Q1 – Q3]. Correlations between sex, age, eGFR, and UCr-normalized AQP2+ or PODXL+ uEV concentrations were examined using Spearman’s correlation. Linear regression analyses were performed between eGFR, log-transformed UCr-normalized AQP2+ uEV, and PODXL+ uEV concentrations. A schematic overview of sample collection, processing, and staining is illustrated in **Supplementary Fig. S2**.

## Results

CD63+ AQP2+ uEVs (**Fig. 1a**) and CD63+ PODXL+ uEVs (**Fig. 1b**) were clearly present in urine compared to the negative controls, including detergent treatment, reagent-containing PBS, and isotype staining (**Fig. 1c** – **1g**). The medians of UCr-normalized CD63+ AQP2+ uEV and CD63+ PODXL+ uEV concentrations were 3.5 [Q1 – Q3: 0.8 – 6.8] × 10^4^ objects/μmol and 3.6 [1.2 – 7.7] × 10^4^ objects/μmol, respectively, with less than 20% of these uEVs measured in detergent-treated or isotype-stained urine (**Fig. 1h**; all *p* values < 0.001). With the presence of detergent, 40 – 90% of objects stained negative for CD63 but positive for AQP2 or PODXL (**Fig. 1c, 1d**). In addition, compared to urine (**Fig. 1a, 1b**), reagent-containing PBS contained 10% of CD63-negative and AQP2/PODXL-positive backgrounds (**Fig. 1e, 1f**), indicating free antibody aggregations rather than biological uEVs. No significant association was observed between CD63+ AQP2+ or PODXL+ uEV concentration and sex, age, or eGFR, but a significant correlation was found between AQP2+ and PODXL+ uEV concentrations (Rho = 0.789, *p* < 0.001; **Fig. 1i**). Regression analyses were used to visualize these correlations, which also demonstrated weak associations between eGFR and AQP2+ uEV concentrations (R^2^ = 0.00, *p* = 0.98; **Fig. 1j**) or between eGFR and PODXL+ uEV concentrations (R^2^ = 0.04, *p* = 0.30; **Fig. 1k**). However, a higher level of PODXL+ uEVs was significantly associated with a higher AQP2+ uEV level (R^2^ = 0.64, *p* < 0.001; **Fig. 1l**). To explore whether AQP2 colocalizes with PODXL on the CD63+ uEV surface, we performed CD63-AQP2-PODXL triple staining. About 50% of the CD63+ uEVs expressing PODXL were triple-positive and expressed AQP2 (**Fig. 1m**). Representative images for AQP2+ PODXL+ CD63+ uEVs are displayed in **Fig. 1n**. The specific colocalization of AQP2 and PODXL on CD63+ uEVs was strengthened by the negative controls and rare AQP2 and PODXL double-positivity measured in CD63-events (**Supplementary Fig. S3**). The median of UCr-normalized triple-positive uEV concentration was 6.4 [4.0 – 14.9] × 10^3^ objects/μmol, significantly higher than in controls with detergent treatment (*p* = 0.004) or isotype staining (*p* < 0.001; N = 10; **Fig. 1o**).

**Fig. 1.**
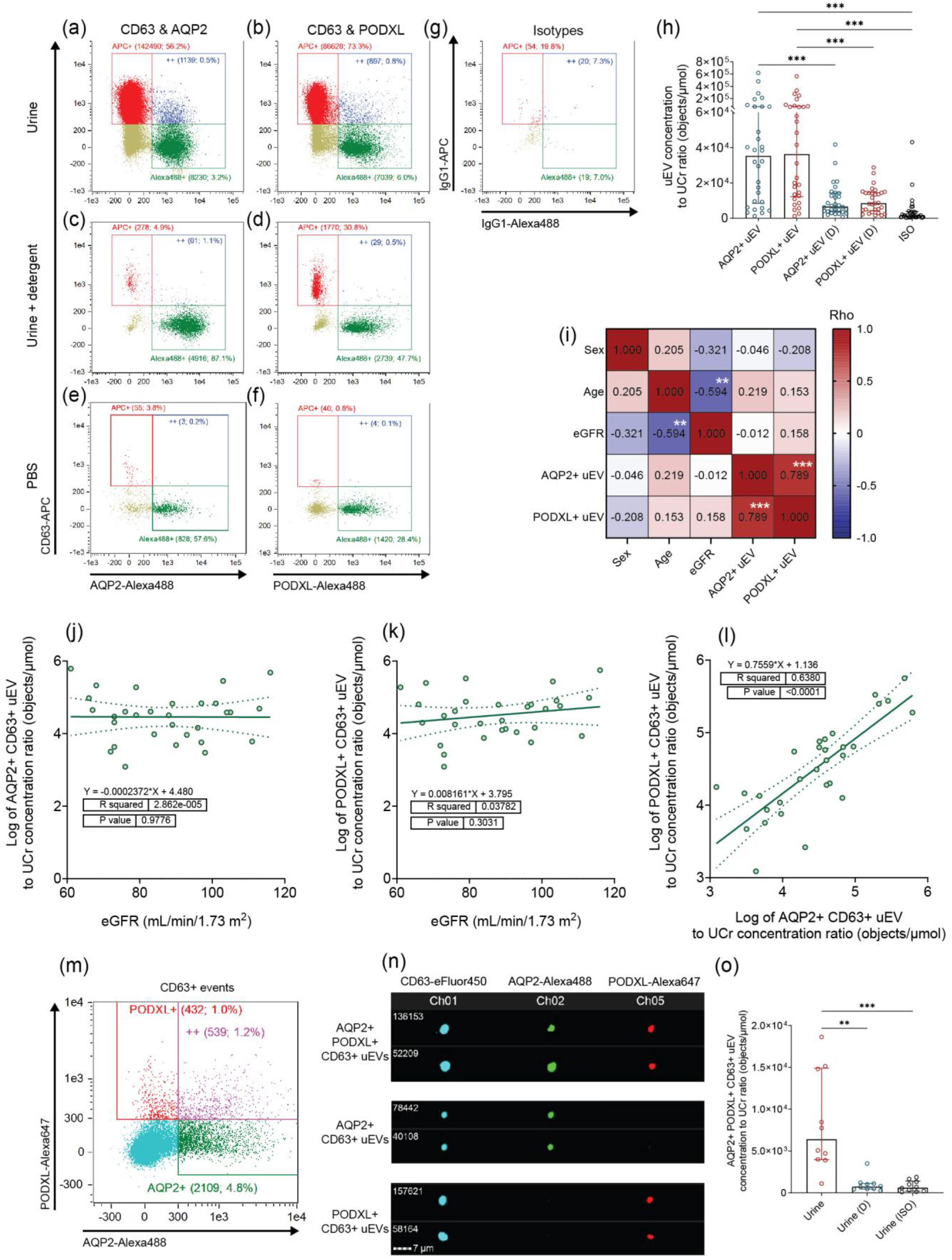
Measurement of kidney-derived urinary extracellular vesicles using IFCM. **a – g** Representative scatterplots of uEV in IFCM. Urine is stained by CD63-APC combined with AQP2-Alexa488 (**a**) or PODXL-Alexa488 (**b**). **c, d** Stained urine was treated with detergent. PBS with CD63-APC and AQP2-Alexa488 (**e**) or PODXL-Alexa488 (**f**). **g** Urine is labeled by IgG-APC and IgG-Alexa488. These scatterplots show CD63-APC single-positives in the “APC+” gate, AQP2 or PODXL-Alexa488 single-positives in “Alexa488+”, and double-positives in “++”. Each gate’s name denotes the count and the percentage of gated objects. **h** A summary of urine samples showing UCr-normalized CD63+ AQP2+ or CD63+ PODXL+ uEV concentrations with controls of detergent treatment and isotype staining. **i** A Spearman test showing correlations among sex, age, eGFR, and UCr normalized kd-uEV levels. The Rho value is presented in each cell to demonstrate the correlation between the corresponding X- and Y-axis parameters. A scale bar of the Rho value is presented on the right of the heatmap. The red and blue colors depict positive and negative correlations, respectively. **j** – **l** Linear regression analyses between eGFR and AQP2+ uEV concentration (**j**), eGFR and PODXL+ uEV concentration (**k**), and between AQP2+ and PODXL+ uEV concentrations (**l**). The concentrations of uEVs are normalized by urine creatinine and log-transferred. **m** The characterization of AQP2 or PODXL positiveness on the CD63+ uEVs. All dots are CD63+. The red gate denotes PODXL positivity, the green gate shows AQP2 expression, and the purple gate double-positive (“++”) signal for AQP2 and PODXL. Each gate’s name includes the gated count and percentage. **n** The representative images for AQP2+ PODXL+ CD63+ uEVs, AQP2+ PODXL-CD63+ uEVs, and PODXL+ AQP2-CD63+ uEVs. The object number is listed on the left of each image. **o** A summary of Ucr-normalized AQP2-PODXL-CD63 triple-positive uEV concentrations measured in urine, urine with detergent treatment, and isotype staining. **Marks:** **, *p* < 0.01; ***, *p* < 0.001. *AQP2* aquaporin 2; *D* detergent-treated control; *eGFR* estimated glomerular filtration rate; *IFCM* imaging flow cytometry; *ISO* isotype-staining control; *PODXL* podocalyxin; *UCr* urine creatinine.

## Discussion

This study investigated the association between kidney biology and podocyte-derived PODXL+ uEVs as well as tubule/collecting duct-derived AQP2+ uEVs. There was no correlation between these kd-uEV levels and sex, age, or eGFR. Despite the substantial anatomical distance between podocytes and collecting duct cells, a significant correlation was found between the uEVs expressing AQP2 and PODXL, which can be explained by the partial colocalization of these proteins on uEVs. Collecting duct cells are active in absorbing podocyte-derived uEVs [7]. Endocytosis is a typical way to uptake external vesicles, allowing the endosomal system to package native molecules and release exosomes, a distinctive uEV subtype marked by CD63 expression [8]. This procedure can lead to vesicle membrane reversion or the presence of inside-out proteins on its surface [9]. Our AQP2 antibody targets its intracellular domain, which typically faces the lumen of vesicles rather than the exterior environment [10]. Therefore, the presence of uEVs with exterior AQP2 further suggests their endosomal origin [10]. Our study provides evidence of EV-mediated protein delivery in the human nephron, which gives new insights into the intra-nephron cross-talk and kidney biology.

The limitation is that other kidney-specific markers on uEVs, such as tubular co-transporters, were not measured. Characterization of more markers can help understand the role of EVs in kidney physiology, but our study, for the first time, demonstrates the colocalization of podocyte and tubular proteins on uEVs. Further investigations are needed to confirm these findings and fully explore uEV-mediated communication in nephron health and disease.

## Conclusion

The release of PODXL+ or AQP2+ CD63+ kd-uEVs is not correlated with kidney function. The correlation between these two uEV populations can be explained by the colocalization of PODXL and AQP2 on CD63+ uEVs.

## Declarations

### Ethical approval

This study including human participants was performed in line with the ethical principles of the Declaration of Helsinki and its later amendments. Approval was granted by the Ethics Committee of Erasmus Medical Center (Ethics approval number: 2018-035).

### Informed consent to participate and publish

Informed consent regarding participating and publishing data was obtained from all individuals included in this study.

### Data availability statement

The data generated during this study are not publicly available due to concerns regarding participant privacy. Data can be accessed from the corresponding author upon reasonable request. The uEV experimental parameters were submitted to the EV-TRACK knowledgebase (ID: EV240051) and are available at the following URL: https://evtrack.org/.

## Acknowledgments

We acknowledge the contribution of Weicheng Xu and Thierry P. P. van den Bosch (EMC) to IFCM and immunohistochemistry experiments. The figure was created on BioRender.com with copyright licenses.

## Supplementary file

This study contains a supplementary file.

## Authors’ contributions

Conceptualization: [Liang Wu], [Carla C. Baan], [Karin Boer]; Methodology: [Liang Wu], [Derek Reijerkerk]; Formal analysis and investigation: [Liang Wu]; Writing - original draft preparation: [Liang Wu]; Writing - review and editing: [Liang Wu], [Carla C. Baan], [Karin Boer], [Derek Reijerkerk], [Dennis A. Hesselink]; Funding acquisition: [Liang Wu], [Carla C. Baan], [Karin Boer]; Resources: [Carla C. Baan], [Karin Boer], [Dennis A. Hesselink]; Supervision: [Carla C. Baan], [Karin Boer], [Dennis A. Hesselink].

