## Supplemental information for "Urinary extracellular vesicles in healthy individuals: positive correlation between podocyte and tubular vesicles independent of kidney function"

### Supplementary Tables and Figures

**Table S1 Clinical characteristics of enrolled subjects (n = 30)**

| Parameters | Values |
|---|---|
| Age (median [Q1 – Q3]; year) | 47.0 [24.0 – 57.5] |
| Sex (male number & percentage) | 16 (53.3%) |
| eGFR (median [Q1 – Q3]; mL/min/1.73 $m^2$)[a] | 86.0 [73.0 – 101.8] |
| BMI (median [Q1 – Q3]; kg/$m^2$) | 26.0 [21.6 – 27.6] |
| UCr concentration (μmol/mL) | 3.4 [1.9 – 6.8] |

[a] eGFR is calculated using the CKD-EPI creatinine equation (2021): Shi J, Lindo EG, Baird GS, et al (2021) Calculating estimated glomerular filtration rate without the race correction factor: Observations at a large academic medical system. Clin Chim Acta 520:16–22. https://doi.org/10.1016/j.cca.2021.05.022.

*AQP2* aquaporin 2; *eGFR* estimated glomerular filtration rate; *PODXL* podocalyxin; *UCr* urine creatinine.

**Supplementary Table S2** Information of antibodies used for EV labeling in IFCM experiments

| Antibody application | Antibody | Manufacturer | Clone | In-stock concentration (µg/mL) | Antibody dilution |
|---|---|---|---|---|---|
| *AQP2+ CD63+ EVs* | CD63-APC | Biolegend ( San Diego, CA, USA) | H5C6 | 200 | 1:30 |
| | AQP2-Alexa488 | Santa Cruz (Dallas, TX, USA) | E-2 | 200 | 1:15 |
| *PODXL+ CD63+ EVs* | CD63-APC | Biolegend | HI9a | 200 | 1:30 |
| | PODXL-Alexa488 | Santa Cruz | 3D3 | 200 | 1:30 |
| *AQP2+ PODXL+ CD63+ EVs* | CD63-eFluor450 | ThermoFisher Scientific (Waltham, MA, USA) | H5C6 | 200 | 1:15 |
| | AQP2-Alexa488 | Santa Cruz | E-2 | 200 | 1:15 |
| | PODXL-Alexa647 | Santa Cruz | 3D3 | 200 | 1:20 |
| *Isotype-staining controls* | IgG1-APC | Biolegend | MOPC-21 | 200 | 1:30 |
| | IgG1-Alexa488 | ThermoFisher Scientific | P3.6.2.8.1 | 200 | 1:15 or 1:30 |
| | IgG1-eFluor450 | ThermoFisher Scientific | P3.6.2.8.1 | 200 | 1:15 |
| | IgG1-Alexa647 | Biolegend | MOPC-21 | 50 | 1:5 |

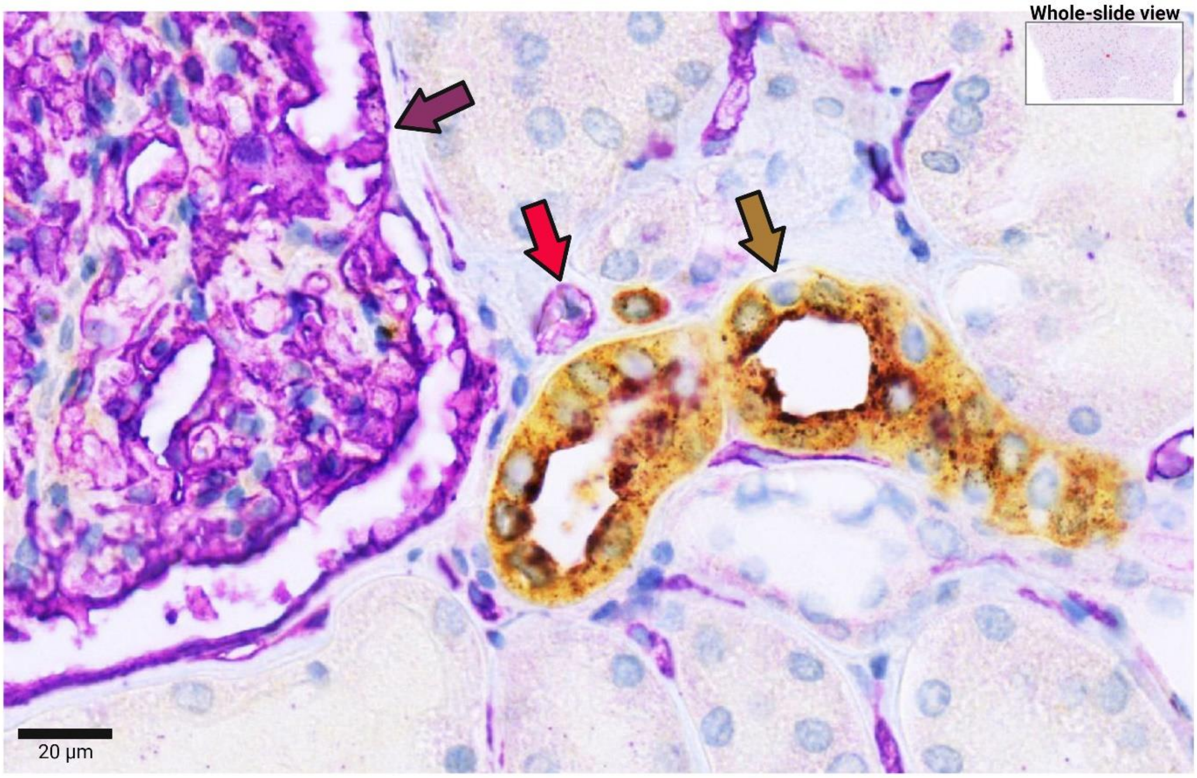


**Supplementary Fig. S1** Immunohistochemistry of peritumoral kidney tissue. The tissue was stained for aquaporin 2 (AQP2; brown) and podocalyxin (PODXL; purple). The brown arrow denotes the collecting duct, and the purple arrow points to the glomerulus demonstrating PODXL-positive podocytes. Peritubular endothelial cells are also slightly positive for PODXL (red arrow).

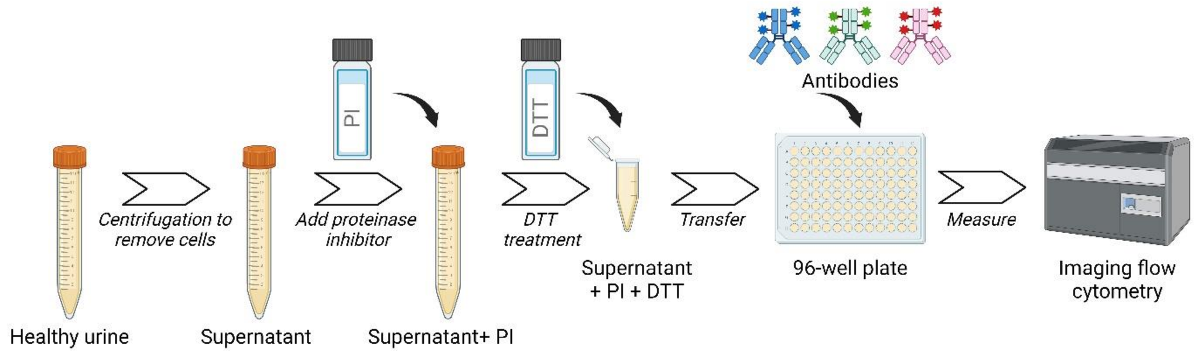


**Supplementary Fig. S2** Schematic overview of urine collection, processing, and labeling for imaging flow cytometry. Created with BioRender.com.

*DTT* dithiothreitol; *PI* proteinase inhibitor.

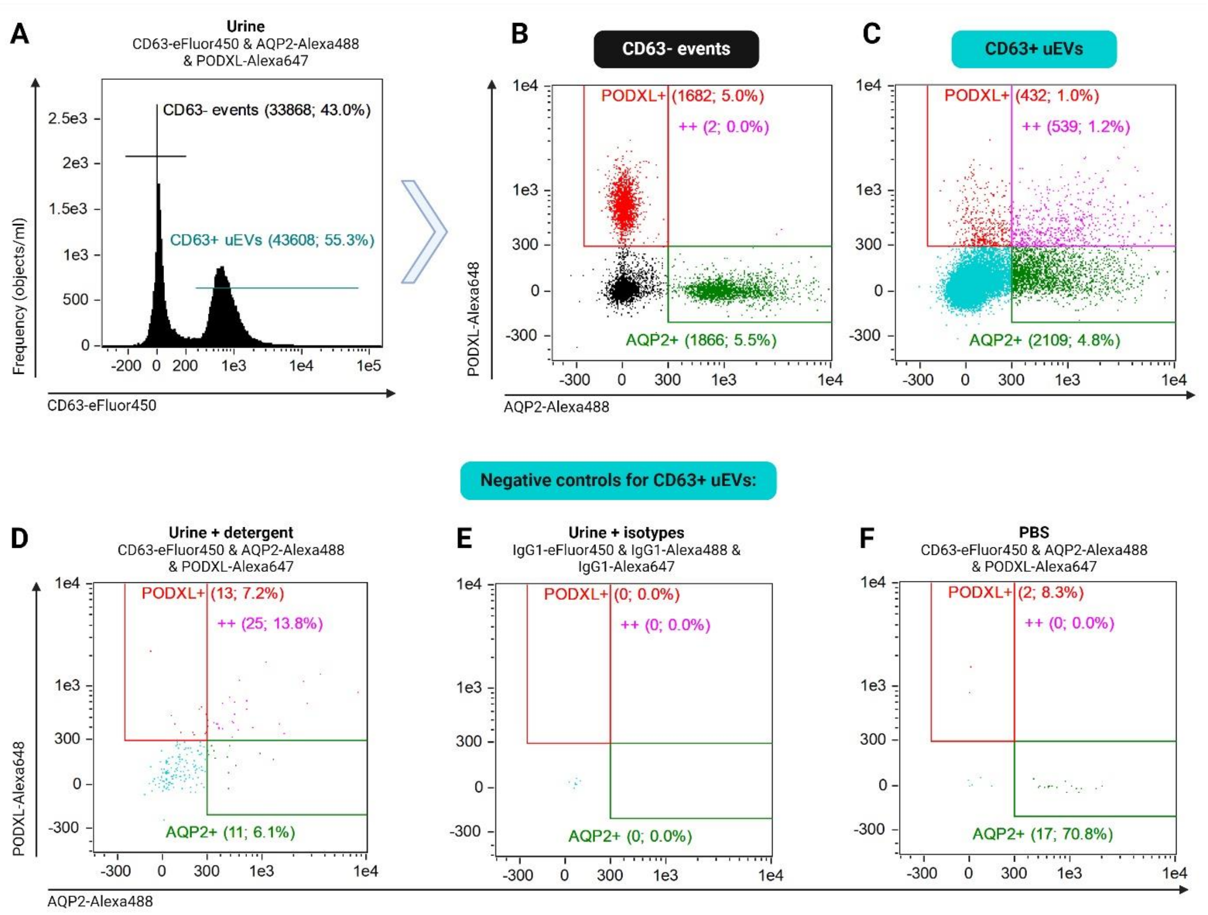


**Supplementary Fig. S3** CD63-AQP2-PODXL triple staining on uEVs using IFCM. **a** A representative scatterplot illustrates the categorization of CD63- events and CD63+ uEVs in accordance with the CD63-eFluor450 intensity. **b**, **c** The positivity of AQP2 or PODXL for CD63- events (**b**) or CD63+ uEVs (**c**). **d** – **e** Representative negative controls validating the triple staining, including CD63-AQP2-PODXL triple-stained urine after detergent treatment (**d**), isotype-stained urine (**e**), and reagent-containing PBS (**f**).
